# Evaluation of a Scalable Design for a Pediatric Telemedicine and Medication Delivery Service: A Prospective Cohort Study in Haiti

**DOI:** 10.1101/2024.08.16.24312128

**Authors:** Molly B. Klarman, Xiaofei Chi, Youseline Cajusma, Katelyn E. Flaherty, Jude Ronald Beausejour, Lerby Exantus, Valery M. Beau de Rochars, Chantale Baril, Torben K. Becker, Matthew J. Gurka, Eric J. Nelson

## Abstract

**Objective:** To evaluate a scalable pediatric telemedicine and medication delivery service (TMDS) that extends early healthcare access at households to avert emergencies.

**Study design:** A TMDS in Haiti was configured for scalability by triaging severe cases to hospital-level care, non-severe cases with higher clinical uncertainty to in-person exams at households with medication delivery, and non-severe cases with low clinical uncertainty to medication delivery alone. This design was evaluated in a prospective cohort study conducted among pediatric patients (≥10 years). Clinical and operational metrics were compared to a formative reference study in which all non-severe patients received an in-person exam. The primary outcomes were reported rates of clinical improvement/recovery and in-person care seeking at 10-days.

**Results:** 1043 cases (41 severe; 1002 non-severe) were enrolled in the scalable TMDS mode and 19% (190) of the non-severe cases received an in-person exam. 382 cases (24 severe, 358 non-severe) were enrolled in the reference study and 94% (338) of the non-severe cases received an in-person exam. At 10-days, rates of improvement were similar for the scalable (97%, 897) and reference (95%, 329) modes. Rates of participants who sought follow-up care were 15% (138) in the scalable mode and 24% (82) in the reference mode.

**Conclusion:** In the context of a five-fold reduction of in-person exams, participants in the scalable TMDS mode had non-inferior rates of improvement at 10-days. These findings highlight an innovative and now scalable solution to improve early access to healthcare at households without compromising safety.

## INTRODUCTION

The World Health Organization (WHO) established the Integrated Management for Childhood Illness (IMCI) guidelines for the management of sick children in 1995 (1, 2). The approach has since been deployed globally and is the current standard for community-based pediatric care in low- and middle-income countries (LMIC) (3). There are numerous challenges to successfully implement the IMCI strategy which limits its potential to effectively reduce morbidity and mortality (4–8). Implementation science frameworks have facilitated the design, evaluation and uptake of potential solutions to address these complex challenges (9). These solutions include technologies aimed at bridging the ‘know-do gap’, specifically the gap between knowledge of the IMCI guidelines and effective implementation of the guidelines (10–14).

Despite these advances, there are gaps in the accessibility of IMCI adherent clinics. We seek to close these gaps by extending early access to high-quality pre-emergency pediatric healthcare at the household level. We focus on the nighttime when healthcare seeking is often delayed to the morning because of financial and logistical barriers (15, 16). Delay raises the risk of developing life-threatening disease that leads to higher-cost interventions (17, 18). In response, we designed and implemented a pediatric telemedicine and medication delivery service (TMDS) that uses clinical resources derived from the IMCI guidelines.

The TMDS model has been developed and rigorously evaluated within the Improving Nighttime Access to Care and Treatment (INACT) study series. The INACT studies began in Haiti (15) and expanded to Ghana to test adaptability (16). In Haiti, the INACT1-H study characterized care-seeking barriers (15). The INACT2-H study was a pre-pilot that established clinical safety, feasibility and guideline performance of a TMDS (19, 20). The study tested if ‘what was heard’ over the phone per virtual exams matched ‘what was seen’ at the bedside per in-person exams. Severe cases were referred to hospital-level care. All non-severe cases received paired virtual and in-person exams to determine the accuracy of the virtual assessments and treatment plans. This study identified high rates of improvement/recovery at 10-days (95%). The evaluation of most non-severe cases had what we considered low clinical uncertainty (see methods), and we hypothesized these cases could safely be managed with medication delivery alone. However, a subset of cases had higher clinical uncertainty, and we hypothesized these cases would require continued in-person evaluation. We reasoned that configuring the TMDS to align with these hypotheses would reduce in-person exams by approximately 85% without compromising the high rates of improvement/recovery.

The INACT3-H study, presented herein, aims to test our clinical guidelines, hypotheses, and overall approach as a pathway to scalability. Within implementation science frameworks for scaling(21), our approach addresses the facilitator; ‘designing the innovation for scale’(22, 23). In doing so, the primary design modification was removing the clinical ‘guardrail’ of in-person exams for non-severe cases with low clinical uncertainty. A secondary design modification was restructuring the TMDS into a ‘hub and spoke’ model where one central call center serviced multiple delivery zones. These modifications are anticipated to reduce the financial and human resources required to operate a TMDS (24). Our approach was respectful of the challenges with real-world implementation of novel innovations and the study protocol was permissive of iterations. The scalable design was implemented within a prospective cohort study (INACT3-H) and the results were compared to the prior published prospective cohort study as a reference standard (INACT2-H).

## METHODS

### Ethics statement

The INACT3-H study and the previously published INACT2-H study were reviewed and approved by the University of Florida Institutional Review Board (IRB202002693; IRB201802920 as well as the Comité National de Bioéthique (National Bioethics Committee of Haiti; Ref2021-11; Ref1819-51).

### Study design

The INACT2-H and INACT3-H studies were prospective cohort studies. The INACT2-H workflow was modified to facilitate scalability; the scalable mode was deployed in the INACT3-H study. Outcomes from INACT3-H were compared to those from INACT2-H (reference standard).

### Study population and setting

INACT2-H was conducted in Gressier, Haiti. INACT3-H was conducted in both Gressier and Les Cayes, Haiti. Gressier has a population of 36,400 and is characterized by rural mountainous regions and more densely populated coastal communities(25). Les Cayes is approximately 175km west of Gressier. It has a population of 152,000 people(25), and over half live in the urban center. On August 14, 2021, Les Cayes was impacted by a 7.2 magnitude earthquake that displaced inhabitants and impacted population dynamics (26). Both Gressier and Les Cayes have minimal public infrastructure (e.g., electricity, paved streets, reliable telecommunication services). The study periods were marked by spikes of political and societal instability that impacted the movement of goods and provision of services with detrimental effects on health and healthcare(27, 28).

### Participant recruitment

Participant populations were informed of the TMDS through radio and print advertisements as well as announcements at schools and clinics.

### Participant inclusion criteria and consent process

For both studies, children were eligible to participate if their parent contacted the TMDS during operational hours (6 PM-5 AM) regarding an illness experienced by their child aged ≥10 years. Written consent from parents/guardians, and assent from children ≥7 years, was obtained from families who received an in-person exam. Parents/guardians whose only contact with the TMDS was a virtual encounter were read a waiver of documentation of consent over the phone.

### Enrollment

Enrollment estimates for the scalable mode (INACT3-H) were based on expected call volume that was established in the reference mode (INACT2-H).

Enrollment during INACT3-H was increased iteratively to assure that both research and service objectives were met in response to dynamic national safety and logistical challenges.

### Participant incentives and fees

Across both studies, TMDS users were asked to pay a fee of 500 Gourdes (5 USD) to contribute to the cost of delivery and medications. The fee was reduced or waived for those who were unable to pay in full.

### Implementation

#### TMDS workflow

The workflow elements shared between the studies were as follows (Figure 1); i) A parent contacted the call center and was connected to a provider. ii) The provider gathered demographic and clinical information about the sick child, referenced clinical resources, and generated a treatment plan that included triage classification (mild, moderate, or severe), treatment location (hospital or home), recommended medications and/or fluids, and advice on follow-up care. iii) Severe cases were immediately referred to the hospital. iv) For non-severe cases, those residing in a delivery zone received an in-person exam by a provider and/or delivery of medications/fluids. v) Children residing outside a delivery zone, or who did not require medications/fluids, received virtual advice alone. vi) Patients requiring hospital or mandatory clinic follow-up received a 24-hour follow-up call. All patients received a 10-day follow-up call to ascertain the child’s condition and feedback on the service.

**Figure 1.**
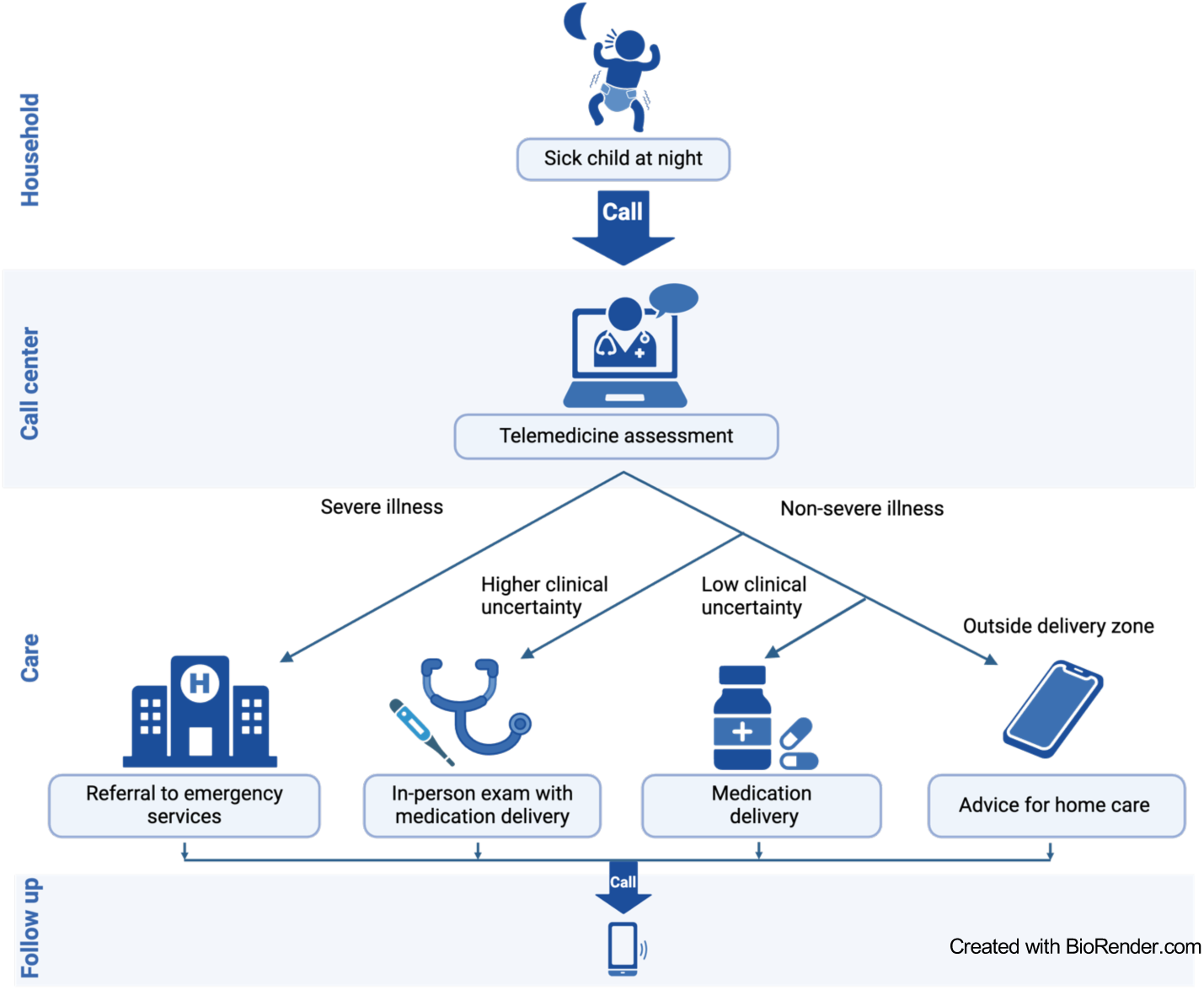
General workflow of a telemedicine and medication delivery service (TMDS). The parent of a sick child contacts the call center. A provider receives the call and performs a virtual exam by asking the parent targeted questions about the child and their specific symptoms. The provider references clinical resources to create a treatment plan based on illness severity that includes disposition, medications/fluids, and follow-up recommendations. Families are contacted by phone at 10 days; severe cases are also contacted at 24 hours. In the reference mode (INACT2H) all non-severe cases residing within the delivery zone received a paired in-person exam at the household to compare evaluate the quality of virtual exams to the in-person exams.

#### TMDS elements specific to the reference mode (INACT2-H)

The call center served a single delivery zone in Gressier, Haiti. Call center providers performed in-person exams for all non-severe cases within the delivery zone; the paired exams were used to assess the accuracy of the virtual exams.

#### TMDS workflow elements specific to the scalable mode (INACT3-H)

The INACT2-H call center and Gressier delivery zone were maintained and a second delivery zone in Les Cayes, Haiti was added in response to humanitarian need caused by an earthquake.

The TMDS workflow was configured to reserve in-person exams for non-severe cases with higher clinical uncertainty. Participant siblings of these cases also received in-person exams. For non-severe cases with low clinical uncertainty, delivery drivers were dispatched to transport medications/fluids to participant households. We estimated this change would limit in-person exams to approximately 15% of all cases. In the ‘delivery-alone’ situations, instructions on how to prepare and administer the medications/fluids were relayed by the provider to the parent over the phone at the end of the virtual exam. The information was reinforced through video tutorials that were played for parents on a tablet at the household; parents were offered an opportunity to reconnect to call center providers for clarifications.

#### Provider and staff training

The providers were licensed Haitian nurses and nurse practitioners. Six of ten call center providers worked for the TMDS during both the INACT2-H and INACT3-H studies. Licensed Haitian physicians were available by phone to provide advice on cases outside the scope of the clinical resources. All providers and drivers received didactic and experiential training relative to their clinical, research, and operational responsibilities.

#### Clinical procedures, guidelines and resources

Providers were equipped with three primary clinical resources to guide the virtual and in-person exams. These resources are described elsewhere (29) and include: (i) A clinical guideline based on in-person WHO pediatric resources and adapted for telemedicine use (Supplementary Text1). The guideline prioritized six common childhood illnesses/problems: ‘Fever’, ‘Respiratory problem/Cough’, ‘Dehydration’/’Vomit’/’Diarrhea’, ‘Ear pain’, ‘Skin problem’, ‘Pain with urination’ and ‘Other’. Each section began with a summary of the problem and was stratified by the diagnostic criteria used to triage the severity of the problem into mild, moderate, or severe categories. Criteria that designated a case as severe were referred to as ‘danger signs’. Within each stratum there was guidance on how to treat, where to treat, and when to follow-up. Directives on treatment location were based on severity and clinical uncertainty. In regard to ‘clinical uncertainty’, a strict definition is difficult to set and build into guidelines because it is the collective impression the provider establishes based on both objective and subjective data. Our guidelines were derived from IMCI guidelines which adhere to broad standards of pediatric care and organically make recommendations based on degrees of uncertainty. With this mindset, our guidelines prioritized in-person exams for patients less than 1 year age, especially those with fever and/or concern for fever and cough, as well as patients with concern for dehydration (e.g., 12+ episodes of diarrhea) or concerns for respiratory insufficiency (e.g., fast breathing). These conditions are associated with high clinical uncertainty as they often present subtly, yet are vulnerable to progressive decompensation, specifically from sepsis, pneumonia, and hypovolemic shock, respectively. (ii) A paper case report form that guided data collection during virtual and in-person exams and was embedded with clinical decision support (Supplementary Text2). (iii) A medication formulary that outlined available medications/fluids and dosing recommendations (Supplementary Text3).

These resources have been iterated and adapted to real-world use since they were first piloted in September 2019. The resources were written for mid-level healthcare providers (e.g., experienced nurses, nurse-practitioners). While the guidelines did not explicitly state when a case scenario has higher versus low clinical uncertainty, uncertainty is built into the algorithms, which follow WHO standards of practice(2, 30). The WHO standards are based in part on risk stratifications set by consensus-based expert opinion or evidence-based primary clinical research studies. The clinical guidelines and resources remain intended for use in this INACT study series alone.

### Outcome measures

#### Primary

The primary outcome measures were rates of improvement/recovery and rates of in-person care seeking at 10-day follow-up.

#### Secondary

Secondary outcome measures consisted of operational, performance and 10-day feedback components. Operational measures were ‘virtual exam duration’ defined as the length of time to complete a virtual exam over the phone and ‘time to arrival at household’ defined as the length of time spanning from initial contact with the TMDS by phone until a driver (with or without provider) arrived at the participant’s house. Performance measures focused on congruence and guideline deviations.

Congruence refers to the agreement between components of the paired virtual and in-person exams; congruence was assessed for case severity, presence of fever, fast breathing, and dehydration, and treatment with select medications. Guideline deviations were instances when a provider made a clinical decision outside the scope of the written clinical guidelines. Deviations were expected and may have occurred due to human error, situational adaptability, or direction from the oversight physician.

Deviations were quantified for danger signs and select medications provided. User feedback was obtained in structured (Likert scale) and unstructured (free text) formats. *Exploratory*. Analyses were stratified by type of care provided and delivery zone location.

### Data analysis

Clinical, operational, and performance outcome measures were compared between the scalable (INACT3-H) and reference (INACT2-H) modes. *Analytic strategy*. Repeat cases within 30 days were excluded from both study cohorts. Participants not reached at the 10-day follow-up were excluded from the clinical outcome and feedback analyses. Missing data for other variables was addressed through pairwise deletion. In the scalable mode guideline deviations were identified by internally reviewing the data for instances of missed danger signs and accuracy of medications included in treatment plans (e.g., complaints of pus in ear should yield inclusion of amoxicillin in plan). The identification of guideline deviations occurring in the reference mode was described previously(19). When calculating guideline deviation rates, the number of completed exams (virtual and in-person) was used as the unit of analysis. Congruence was assessed for the subset of cases that had paired virtual and in-person exams which mainly included cases with higher clinical uncertainty.

Comparisons that involved a guideline deviation were excluded from the respective congruence analyses. Medications included in the congruence analyses were selected because of their centrality to the clinical guidelines and were provided most frequently. *Statistical analysis*. Proportions were used to describe categorical variables and medians with interquartile ranges were used to describe continuous variables. To test differences between the modes (scalable and reference) or between locations within the scalable mode (Gressier and Les Cayes), Chi-square tests or Fisher’s exact tests were used for nominal variables, Cochran-Armitage Trend tests for ordinal variables, and two-sample t-tests for continuous variables. To compare clinical and operational outcomes between the two modes stratified by type of care provided, Cochran-Mantel-Haenszel tests (General Association) were used for nominal variables, Cochran-Mantel-Haenszel tests (Row Mean Scores Differ) for ordinal variables, and two-way ANOVA for continuous variables. Binary assessments were described using Cohen’s kappa, sensitivity, specificity, positive predictive values (PPV), and negative predictive values (NPV). Assessments with kappa values were classified as follows: - 0.1 – 0.20 ‘no agreement’, 0.21 – 0.39 ‘minimal’, 0.40 – 0.59 ‘weak’, 0.60 – 0.79 ‘moderate’, 0.80 – 0.90 ‘strong’, and >0.90 ‘almost perfect’(31). Analyses were completed using Statistical Analysis Software (SAS Institute) V.9.4.

## RESULTS

### Participant and case characteristics

The reference mode enrolled 391 participants from September 9, 2019, to January 19, 2021 and the scalable mode enrolled 1068 participants from January 20, 2021 to September 21, 2022. Among enrolled participants, 1043 from the scalable mode and 382 from the reference mode were included in the analyses (Figure 2). The median age was 24 months (IQR 12-48 scalable; 9-48 reference;) for both cohorts, sex distributions were similar (Table 1). There was a lower proportion of participants aged <2 months in the scalable cohort (1%, 15) compared to the reference cohort (7%, 26). The chief and general complaints were similar among both modes. The most prevalent chief complaint was ‘fever’ (33%, 342 scalable; 42%, 159 reference) followed by ‘respiratory problem’ (24%, 247 scalable; 17%, 67 reference), whereas the most prevalent general complaint was ‘respiratory problem’ (66%, 686 scalable; 65%, 247 reference), followed by ‘fever’ (55%, 576 scalable; 55%, 212 reference). Virtual complaints of ‘fever’ were determined subjectively in all but 7 cases (1, 1% scalable; 6, 3% reference). Gastrointestinal and dermatologic complaints were also common. Based on the virtual exams, approximately three-fourths of cases were triaged as mild in both the scalable (76%, 788) and reference (73%, 278) modes. In the scalable mode, 18% (190) of cases received a virtual exam and an in-person exam with delivery and 73% (760) received a virtual exam with delivery. In the reference mode, 88% (338) of cases received a virtual and an in-person exam with delivery. The proportion of cases that received a virtual exam alone (no in-person exam or delivery) because of hospital referral or workflow constraint (e.g., caller was outside delivery zone) was similar between the scalable (9%, 93) and reference (12%, 44) modes.

**Figure 2.**
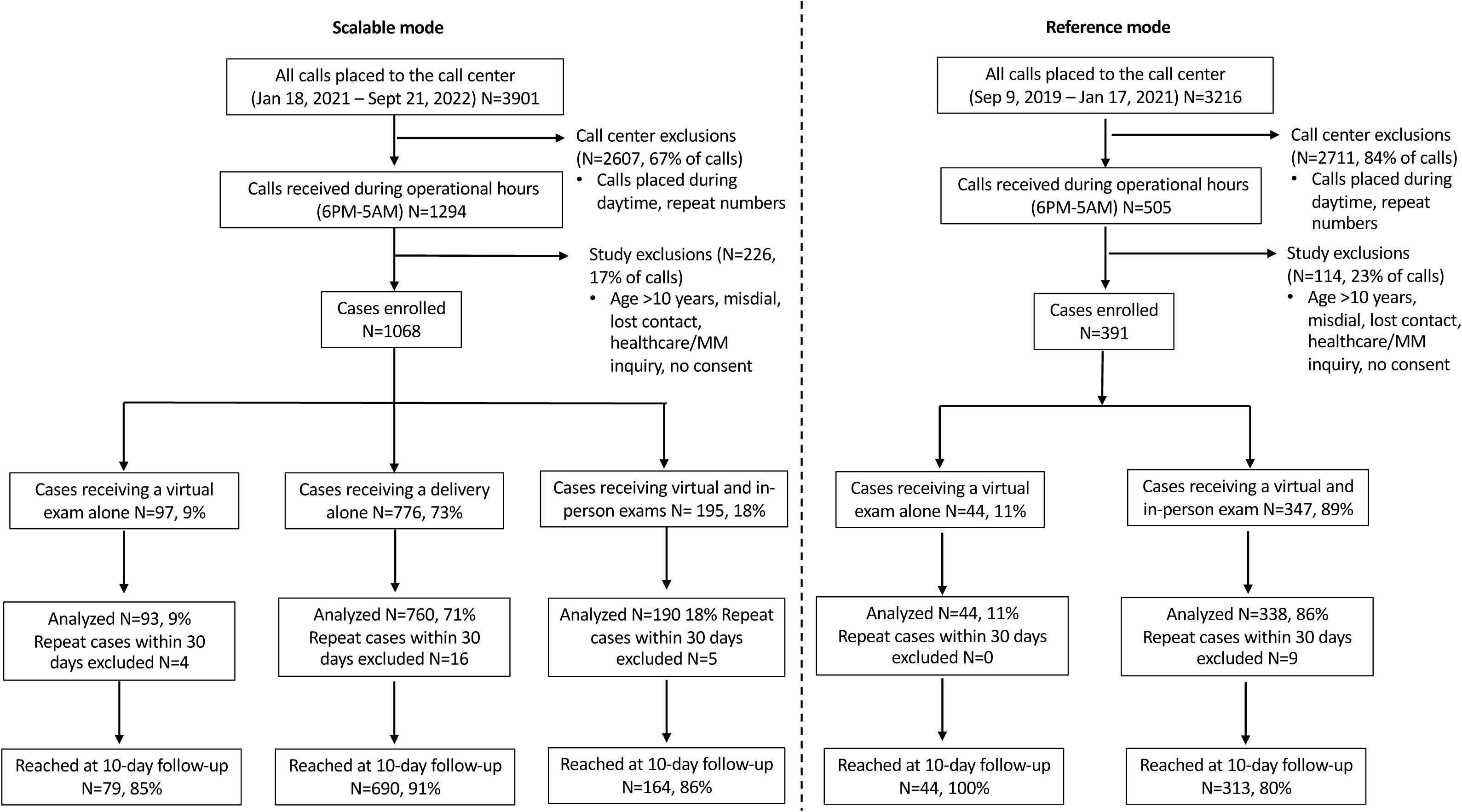
Diagram of case enrollment and inclusion in analyses for scalable and reference Telemedicine and Medication Delivery Service (TMDS) modes.

**Table 1.** Characteristics of participants and cases enrolled in the scalable (INACT3-H) and reference (INACT2-H) modes.

| Characteristic | Scalable mode<br>(n=1043)<sup>a</sup> | Reference mode<br>(n=382)<sup>b</sup> |
| --- | --- | --- |
| Participant age; median mos (Q1-Q3)<sup>c</sup> | 24 (12-48) | 24 (9-48) |
| <2 mos | 15 (1%) | 26 (7%) |
| 2 mos to <2 yrs | 417 (40%) | 164 (43%) |
| 2 yrs to <5 yrs | 374 (36%) | 118 (31%) |
| 5 yrs to 10 yrs | 237 (23%) | 74 (19%) |
| Participant sex; female | 496 (48%) | 178 (47%) |
| Participant chief complaint<sup>d</sup> |  |  |
| Fever | 342 (33%) | 159 (42%) |
| Respiratory | 247 (24%) | 67 (17%) |
| Gastrointestinal | 109 (10%) | 58 (15%) |
| Dermatologic | 237 (23%) | 56 (15%) |
| Other | 108 (10%) | 42 (11%) |
| Participant complaints general<sup>d,e</sup> |  |  |
| Fever | 576 (55%) | 212 (55%) |
| Respiratory | 686 (66%) | 247 (65%) |
| Gastrointestinal | 337 (32%) | 137 (36%) |
| Dermatologic | 383 (37%) | 100 (26%) |
| Case severity at call center |  |  |
| Mild | 788 (76%) | 278 (73%) |
| Moderate | 214 (21%) | 80 (21%) |
| Severe | 41 (4%) | 24 (6%) |
| Type of care provided |  |  |
| Virtual exam and in-person exam with delivery | 190<sup>f</sup> (18%) | 338 (88%) |
| Virtual exam with delivery | 760 (73%) | 0 |
| Virtual exam alone | 93 (9%) | 44 (12%) |

<sup>a</sup> 25 repeat patients within 30 days were excluded.
<sup>b</sup> 9 repeat patients within 30 days were excluded.
<sup>c</sup> (Q1-Q3) = Quartile 1 to quartile 3.
<sup>d</sup> The four most common complaints are represented.
<sup>e</sup> Multiple complaints per participant were permitted.
<sup>f</sup> 19 mild and 3 moderate cases were triaged for delivery but received in-person exams.

### Primary outcomes

In the scalable mode, the rate of participants with a clinical status of improved/recovered was 97% (897) and in the reference mode the rate was 95% (329). No mortalities occurred in the scalable mode, and one occurred in the reference mode. At the 10-day follow up 15% (138) of participants in the scalable mode had sought care and 24% (82) of participants in the reference mode had sought care (Table 2).

**Table 2.**
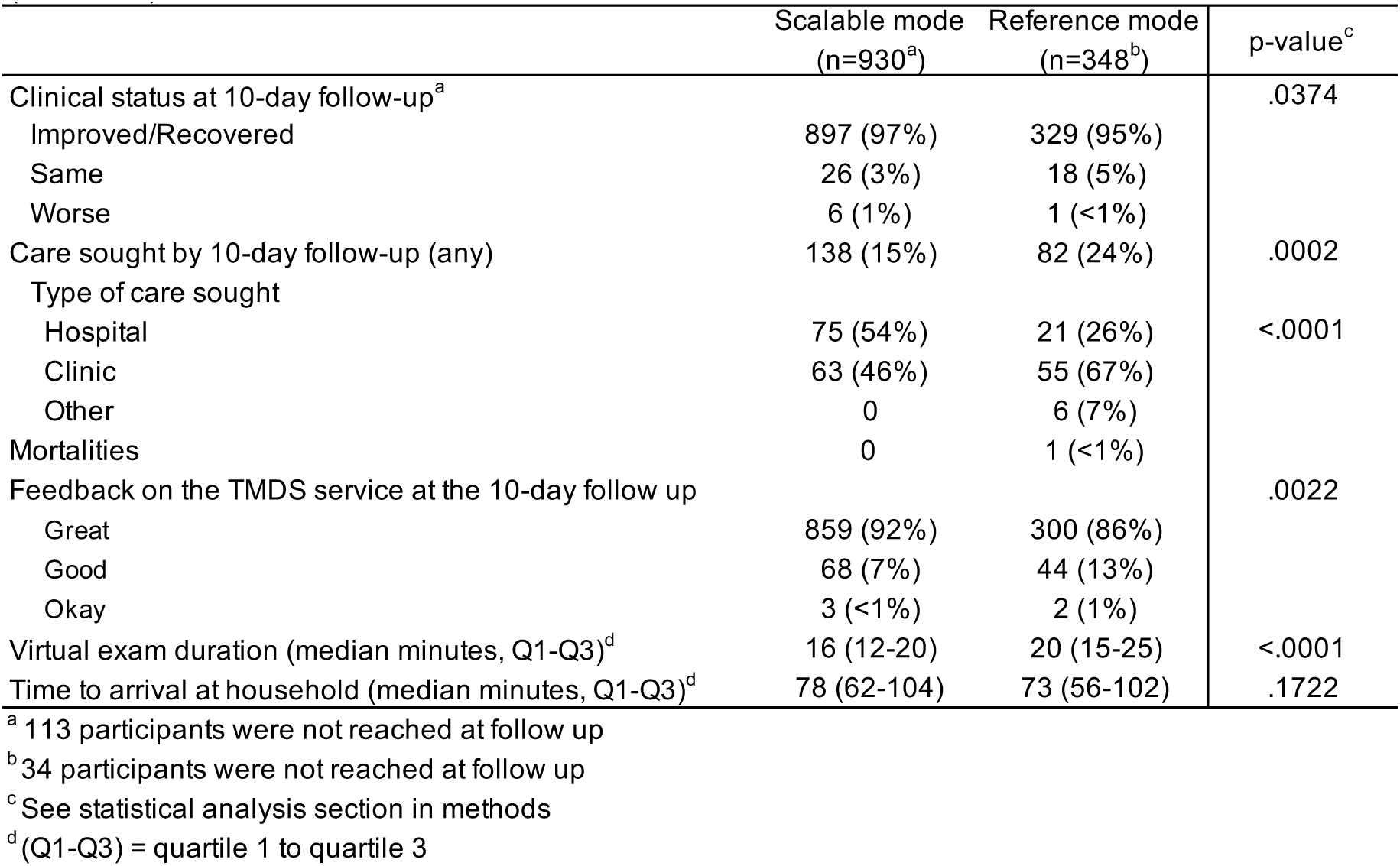
Comparison of clinical and operational outcomes between the scalable (INACT3-H) and reference (INACT2-H) modes.

### Secondary outcomes

The median call duration was 16 mins (IQR 12-20 mins) in the scalable mode and 20 mins (IQR 15-25 mins) in the reference mode. The median time to household arrival was 73 mins (IQR 56-102 mins) in the scalable mode and 78 mins (IQR 62-104 mins) in the reference mode.

The aggregate rate of guideline deviations was 5% (63) in the scalable mode and 11% (82) in the reference mode (Table 3). Rates of missed danger signs were 1% (11) in the scalable mode compared to 2% in the reference mode; a misinterpreted respiratory rate was the most frequent danger sign deviation. Events of medications provided (most commonly amoxicillin) that were not indicated occurred in 1% (18) of cases in the scalable mode and 3% (19) in the reference mode. Events of indicated medications (most commonly zinc) that were not provided occurred in 3% (34) of cases treated in the scalable mode and 7% (52) in the reference mode.

**Table 3.** Comparison of guideline deviations between the scalable (INACT3-H) and reference (INACT2-H) modes.

| Type of guideline deviation | Scalable mode | Reference mode | p-value |
| --- | --- | --- | --- |
| Total deviations (all types) | 63 (5%) | 82 (11%) | <.0001 |
| Danger sign present but not identified (any) | 11 (1%) | 11 (2%) | .1991 |
| General WHO<sup>b</sup> | 0 | 1 (<1%) | .0262 |
| Fever (<2 yrs) | 0 | 2 (<1%) |  |
| Respiration rate | 4 (<1%) | 6 (1%) |  |
| Heart rate | 7 (1%) | 1 (<1%) |  |
| Oxygen saturation<sup>c</sup> | 0 | 1 (<1%) |  |
| Provided medication<sup>d</sup> when not indicated (any) | 18 (1%) | 19 (3%) | .0652 |
| Amoxicillin | 9 (1%) | 17 (2%) | .0008 |
| Benzyl benzoate | 0 | 0 |  |
| Cephalexin | 0 | 0 |  |
| Doxycycline | 0 | 0 |  |
| Erythromycin | 0 | 2 (<1%) |  |
| Paracetamol | 7 (<1%) | 0 |  |
| Zinc | 2 (<1%) | 0 |  |
| Did not provide medication<sup>d</sup> when indicated (any) | 34 (3%) | 52 (7%) | <.0001 |
| Amoxicillin | 0 | 2 (<1%) | .0095 |
| Benzyl benzoate | 2 (<1%) | 0 |  |
| Cephalexin | 0 | 0 |  |
| Doxycycline | 0 | 0 |  |
| Erythromycin | 3 (<1%) | 0 |  |
| Paracetamol | 0 | 0 |  |
| Zinc | 29 (3%) | 50 (7%) |  |

<sup>a</sup> Each virtual and in-person encounter is represented independently.
<sup>b</sup> Lethargic, convulsions, unable to drink/breastfeed
<sup>c</sup> Oxygen saturation was evaluated during in-person exams only.
<sup>d</sup> The most frequently used medications are included.

In the scalable mode congruence was evaluated for the subset of cases that had paired virtual and in-person exams. In this context, the Cohen’s kappa value for cases virtually triaged as mild was minimal at 0.22 (95% CI 0.07-0.38) (Table S1); in the reference mode it was 0.78 (95% CI 0.69-0.87). The kappa value for cases virtually triaged as moderate was 0.25 (0.10-0.41); in the reference mode it was 0.78 (95% CI 0.69-0.87). A similar approach was taken to evaluate select components of the virtual exam. The assessment of fever had a kappa value of 0.24 (95%CI 0.12-0.35); in the reference mode, the corresponding kappa value was 0.58 (95%CI 0.49-0.66). The assessment of fast breathing was not congruent with measured rates during the in-person exam; minimal congruence was observed in the reference mode. The assessment of dehydration during the virtual exam had minimal agreement (kappa 0.24; 95%CI 0.01-0.47); in the reference mode there was moderate agreement (kappa 0.69; 95%CI 0.41-0.98). Despite low congruence, aspects of the virtual exam had high performance metrics (sensitivity, specificity, PPV, NPV) of over 90%. Agreement between medication recommendations based on paired virtual and in-person exams was also assessed. In the scalable mode, paracetamol and amoxicillin exhibited minimal (kappa 0.35; 95%CI 0.22-0.48) and weak (0.47 95% CI 0.35-0.58) agreement, respectively. Benzyl benzoate (0.66 95%CI 0.49-0.83), cephalexin (0.62 95%CI 0.38-0.85) and zinc (0.62 95%CI 0.48-0.75) exhibited moderate congruence. These measures were consistently lower in the scalable mode than the reference mode.

#### Feedback

Participants from the scalable mode rated the service as ‘great’ (highest Likert scale response) at a rate of 92% (859) compared to 86% (300) in the reference mode (Table 2).

### Exploratory

Stratified analyses were performed for both the scalable and reference modes as an exploratory step to evaluate how the type of care provided (virtual exam alone; virtual exam and in-person exam with delivery; virtual exam with delivery) impacted primary and secondary outcome measures. Within both modes, participants who received virtual exams with delivery (with or without in-person exams) had similar rates of improvement/recovery, care seeking, and feedback assessed as ‘great’. The relatively few cases in both the scalable and reference modes who received virtual exams alone without delivery had less favorable outcomes for clinical status, care-seeking and feedback at 10-days (Table 4). Given that the scalable mode included a second delivery zone, we conducted analyses to compare outcome measures between the two sites. Clinical status, rates of care seeking and feedback at the 10-day follow-up were similar between the delivery zones (Table S2). However, compared to Gressier, cases in Les Cayes more commonly sought follow-up care from a hospital and the median time to household arrival was longer.

**Table 4.**
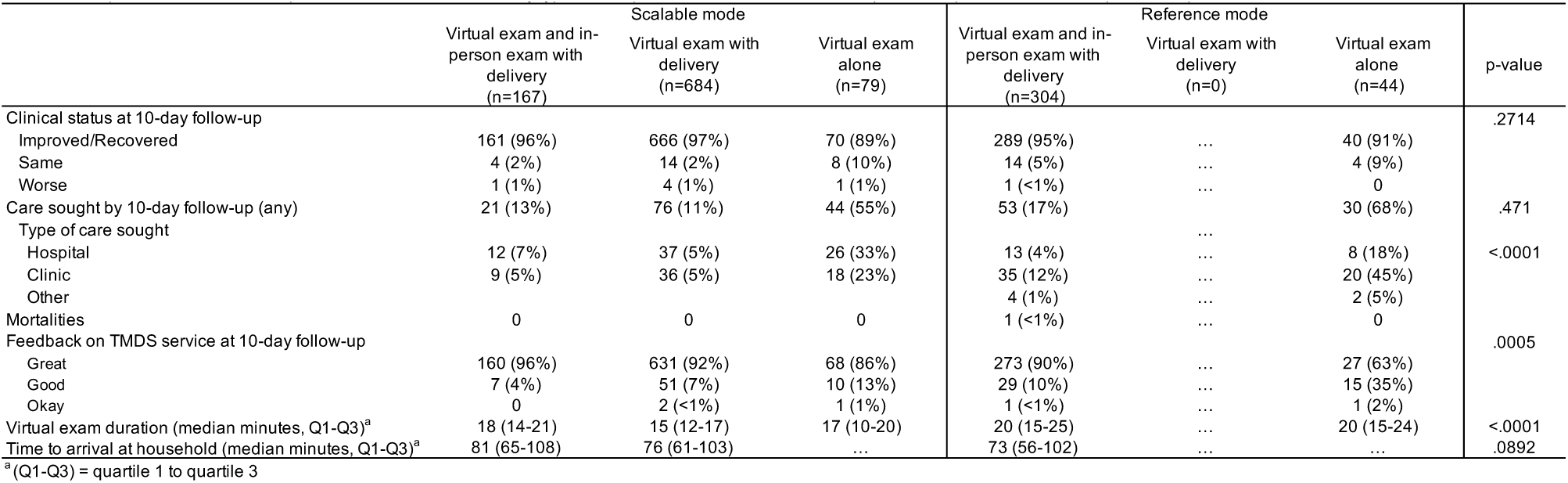
Comparison of clinical and operational outcomes stratified by type of care provided between the scalable (INACT3-H) and the reference (INACT2-H) modes.

## DISCUSSION

We compared a scalable mode of a TMDS that reserved in-person exams for non-severe cases with higher clinical uncertainty to a reference mode that required in-person exams for all non-severe cases; severe cases in both modes were referred to hospital-level care. Pediatric patients were prospectively enrolled in two cohort studies at different time periods to evaluate and compare the two modes. The primary outcome measures were clinical status and rate of in-person care seeking at 10-days. In the context of a five-fold reduction of in-person exams between the scalable and reference modes, rates of improvement/recovery and in-person care seeking at 10-days were similar for the scalable mode. These clinical findings, complemented by similar operational and feedback metrics, represent a critical step towards a scalable solution to improve early access to pediatric healthcare without compromising safety.

The implementation of the INACT studies in Haiti required an adaptative approach to real-world events (e.g., natural disasters, political collapse, insecurity) that in some ways increased the generalizability of the study series to other communities faced with similar challenges. Safety of patients, providers and research staff were prioritized at all times. With this mindset, the design of the reference study required in-person exams at households for all non-severe cases as a clinical ‘guardrail’. This requirement enabled the reference study (INACT2-H) to focus on feasibility, safety, and guideline development/validation(19, 29). In this context we used congruence analyses to evaluate the paired virtual and in-person exams. The analyses exposed anticipated limitations with virtual exams that might leave providers with higher or lower clinical uncertainty depending on the clinical scenario. The clinical guidelines were developed to accommodate for this challenge. For the INACT3-H study, we hypothesized that safety would not be compromised if the ‘guardrail’ of in-person exams was removed for cases with low clinical uncertainty and maintained for cases with higher clinical uncertainty. The results support this hypothesis, thus marking a critical step towards scalability.

Among cases with higher clinical uncertainty that required in-person exams in the scalable mode, there was minimal/weak congruence between the virtual and in-person exams. This finding contrasts the reference study where congruence was higher. The difference was expected as the paired cases in the scalable mode had a higher degree of clinical uncertainty than those in the reference mode by design. Taken together the results support the need for continued in-person exams for cases with higher clinical uncertainty. Exploratory analyses characterized the subset of participants who received a virtual exam with delivery because this group received less contact with a provider compared to those that received a virtual exam and an in-person exam with delivery. Similar clinical outcomes and feedback scores were observed between these two groups. These data suggest a non-inferior patient experience for those cases that received virtual exams with delivery. The small subset of participants who received a virtual exam alone (no in-person encounter by provider or delivery driver) may have had inferior outcomes in terms of clinical status and feedback at 10-days. This finding is not unexpected considering these participants received advice only without provided medication.

One impactful ‘real-world’ event was an earthquake that occurred southwest of the Gressier site soon after initiating the scalable mode (32). For humanitarian reasons, a delivery zone in Les Cayes was added which required an additional step of relaying virtual treatment plans from the Gressier call center to on-call providers in Les Cayes. This adaptation could represent a potential study limitation, however stratification by study site yielded similar outcomes between the two delivery zones. The rate of care seeking at the hospital level was higher in Les Cayes, likely due to increased hospital access within the delivery zone. Les Cayes participants also experienced a longer median time to household arrival that was likely caused by to the need to mobilize the on-call provider; the road network in Les Cayes is better and may have mitigated the extent of the longer arrival time.

The results from this study should be viewed within the context of the following additional limitations: (i) Virtual treatment plans only addressed complaints that parents chose to report. In contrast, in-person treatment plans addressed all problems identified by the provider, allowing virtual and in-person plans to be based on different problems. This factor limited the treatment plan congruence analyses. ii) The clinical resources have undergone minor iterations throughout both study periods. Therefore, the treatment plans were not absolutely equivalent at all points. In addition, we did not control for increased provider expertise over the study period. iii) The environment in which both studies were conducted was volatile which may have confounded enrollment, operational outcomes, and disease severity over time. iv) The primary outcome measures were based on parent reports from the 10-day follow-up call and therefore is at risk of response and recall bias. Despite these limitations, the study provides evidence to justify a transition to the scalable mode.

## CONCLUSIONS

Early access to healthcare is essential to avert morbidity and mortality. However, innovative strategies are needed to assure early access to care, especially when patients are isolated by geography, poverty or illness that presents ‘off-hours’. A telemedicine and medication delivery service (TMDS) represents an innovative solution to extend care to these vulnerable populations. We evaluated a scalable design for a pediatric TMDS for low-resource settings that triaged severe cases to hospital-level care, non-severe cases with higher clinical uncertainty to in-person exams at households with medication delivery, and non-severe cases with low clinical uncertainty to medication delivery alone. We found that within the context of a five-fold reduction of in-person exams, participants in the scalable mode had non-inferior rates of improvement/recovery and care-seeking. Future steps towards scaling will be to digitize the TMDS clinical resources, implement this digital tool within the scalable TMDS workflow and evaluate outcomes.

## Data Availability

De-identified data from this study are available from the corresponding author (EJN) upon request.

## Acknowledgements

We would like to thank both the INACT3-H and INACT2-H study participants and the TMDS employees. We are grateful to the University of Florida Department of Pediatrics administrators, Randy Autrey, Krista Berquist and Brittney Johnson. We express our appreciation for the academic support provided by Glenn Morris at the Emerging Pathogens Institute and Rashmin Savani Chair of the Department of Pediatrics. Finally, we value our partnership with the Ministry of Public Health and Population (Ministère de la Santé Publique et de la Population - MSPP).

## Notes

### Competing Interest Statement

The authors have declared no competing interest.

### Funding Statement

This work was financially supported at the University of Florida by the Children's Miracle Network (EJN), private donations made to the University of Florida Foundation, and a National Institutes of Health grant [5R21TW012332-02] (EJN).

### Author Declarations

The University of Florida Institutional Review Board gave ethical approval for this work (IRB202002693; IRB201802920. Additionally the Comité National de Bioéthique in Port-au-Prince Haiti gave ethical approval for this work (Ref2021-11; Ref1819-51).

### Summary of Updates

One of the funding sources was clarified by adding the grant number.

## REFERENCES

1. Gove S. Integrated management of childhood illness by outpatient health workers: technical basis and overview. The WHO Working Group on Guidelines for Integrated Management of the Sick Child. Bull World Health Organ. 1997;75 Suppl 1(Suppl 1):7–24.

2. World Health Organization. Handbook: IMCI Integrated Management of Childhood Illness Geneva Switzerland2000 [Available from: https://apps.who.int/iris/bitstream/handle/10665/104772/9789241506823_Module-1_eng.pdf?sequence=3.

3. Boschi-Pinto C, Labadie G, Dilip TR, Oliphant N, Dalglish SL, Aboubaker S, et al. Global implementation survey of Integrated Management of Childhood Illness (IMCI): 20 years on. BMJ Open. 2018;8(7):e019079.

4. Kiplagat A, Musto R, Mwizamholya D, Morona D. Factors influencing the implementation of integrated management of childhood illness (IMCI) by healthcare workers at public health centers & dispensaries in Mwanza, Tanzania. BMC Public Health. 2014;14:277.

5. Kim K, Helena H, Albert D, Beatiwel Z, Lumbani B, Josephine L, et al. Integrated Management of Childhood Illnesses (IMCI): a mixed-methods study on implementation, knowledge and resource availability in Malawi. BMJ Paediatrics Open. 2021;5(1):e001044.

6. Bryce J, Victora CG, Habicht JP, Black RE, Scherpbier RW. Programmatic pathways to child survival: results of a multi-country evaluation of Integrated Management of Childhood Illness. Health Policy Plan. 2005;20 Suppl 1:i5–i17.

7. Carai S, Kuttumuratova A, Boderscova L, Khachatryan H, Lejnev I, Monolbaev K, et al. Review of Integrated Management of Childhood Illness (IMCI) in 16 countries in Central Asia and Europe: implications for primary healthcare in the era of universal health coverage. (1468-2044 (Electronic)).

8. Reñosa MA-O, Dalglish SA-O, Bärnighausen KA-O, McMahon S. Key challenges of health care workers in implementing the integrated management of childhood illnesses (IMCI) program: a scoping review. (1654-9880 (Electronic)).

9. Bauer MS, Kirchner J. Implementation science: What is it and why should I care? Psychiatry Research. 2020;283:112376.

10. Sarrassat S, Lewis JJ, Some AS, Somda S, Cousens S, Blanchet K. An Integrated eDiagnosis Approach (IeDA) versus standard IMCI for assessing and managing childhood illness in Burkina Faso: a stepped-wedge cluster randomised trial. BMC Health Serv Res. 2021;21(1):354.

11. Mitchell M, Hedt-Gauthier BL, Msellemu D, Nkaka M, Lesh N. Using electronic technology to improve clinical care – results from a before-after cluster trial to evaluate assessment and classification of sick children according to Integrated Management of Childhood Illness (IMCI) protocol in Tanzania. BMC Medical Informatics and Decision Making. 2013;13(1):95.

12. Schmitz T, Beynon F, Musard C, Kwiatkowski M, Landi M, Ishaya D, et al. Effectiveness of an electronic clinical decision support system in improving the management of childhood illness in primary care in rural Nigeria: an observational study. BMJ Open. 2022;12(7):e055315.

13. Beynon F, Guérin F, Lampariello R, Schmitz T, Tan R, Ratanaprayul N, et al. Digitalizing Clinical Guidelines: Experiences in the Development of Clinical Decision Support Algorithms for Management of Childhood Illness in Resource-Constrained Settings. Global Health: Science and Practice. 2023;11(4):e2200439.

14. Jensen C, McKerrow NA-O, Wills GA-OX. Acceptability and uptake of an electronic decision-making tool to support the implementation of IMCI in primary healthcare facilities in KwaZulu-Natal, South Africa. (2046-9055 (Electronic)).

15. Klarman M, Schon J, Cajusma Y, Maples S, Beau de Rochars VEM, Baril C, et al. Opportunities to catalyse improved healthcare access in pluralistic systems: a cross-sectional study in Haiti. BMJ Open. 2021;11(11):e047367.

16. Flaherty KE, Klarman MB, Zakariah AN, Mahama MN, Osei-Ampofo M, Nelson EJ, et al. Evaluating the prerequisites for adapting a paediatric nighttime telemedicine and medication delivery service to a setting with high malarial burden: A cross-sectional pre-implementation study. Trop Med Int Health. 2023;28(9):763–70.

17. Update: outbreak of cholera ---Haiti, 2010. (1545-861X (Electronic)).

18. Sack DA, Sack Rb Fau - Nair GB, Nair Gb Fau - Siddique AK, Siddique AK. Cholera. (1474-547X (Electronic)).

19. Klarman MB, Flaherty KE, Chi X, Cajusma Y, Capois AC, Vladimir Dofiné MD, et al. Implementation of a pediatric telemedicine and medication delivery service in a resource-limited setting: A pilot study for clinical safety and feasibility. The Journal of Pediatrics. 2022.

20. Molly BK, Xiaofei C, Youseline C, Katelyn EF, Anne Carine C, Michel Daryl Vladimir D, et al. Development and Evaluation of a Clinical Guideline for a Pediatric Telemedicine and Medication Delivery Service: A Prospective Cohort Study in Haiti. medRxiv. 2023:2023.02.15.23285858.

21. Trisha G, Chrysanthi P. Spreading and scaling up innovation and improvement. BMJ. 2019;365:l2068.

22. Spicer N, Hamza YA, Berhanu D, Gautham M, Schellenberg J, Tadesse F, et al. ‘The development sector is a graveyard of pilot projects!’ Six critical actions for externally funded implementers to foster scale-up of maternal and newborn health innovations in low and middle-income countries. Globalization and Health. 2018;14(1):74.

23. Spicer N, Bhattacharya D, Dimka R, Fanta F, Mangham-Jefferies L, Schellenberg J, et al. ‘Scaling-up is a craft not a science’: Catalysing scale-up of health innovations in Ethiopia, India and Nigeria. Social Science & Medicine. 2014;121:30–8.

24. Flaherty KE, Klarman MB, Cajusma Y, Schon J, Exantus L, Beau de Rochars VM, et al. A Nighttime Telemedicine and Medication Delivery Service to Avert Pediatric Emergencies in Haiti: An Exploratory Cost-Effectiveness Analysis. The American Journal of Tropical Medicine and Hygiene. 2022;106(4):1063–71.

25. Direction Des Statistiques Démographiques Et Sociales. Population Totale, De 18 Ans Et Plus. Menages Et Densities Estimes En 2015.; 2015.

26. Eberle C. Technical Report: Haiti earthquake 2021/2022: United Nations University - Institute for Environment and Human Security (UNU-EHS); 2022 [Available from: http://collections.unu.edu/view/UNU:9022#viewMetadata.

27. Taylor L. Haiti’s hospitals plan to close amid fuel shortages. Bmj. 2022;379:o2379.

28. Severe K, Alcenat N, Rouzier V. Resurgence of Cholera in Haiti amidst Humanitarian Crises. N Engl J Med. 2022.

29. Klarman MB, Chi X, Cajusma Y, Flaherty KE, Capois AC, Dofiné MDV, et al. Development and Evaluation of a Clinical Guideline for a Pediatric Telemedicine and Medication Delivery Service: A Prospective Cohort Study in Haiti. medRxiv. 2023:2023.02.15.23285858.

30. World Health Organization. Pocket book of hospital care for children: Second edition Geneva, Switzerland2013 [Available from: https://www.who.int/publications/i/item/978-92-4-154837-3.

31. McHugh ML. Interrater reliability: the kappa statistic. Biochem Med (Zagreb). 2012;22(3):276–82.

32. Daniels JP. Earthquake compounds Haiti’s health challenges. The Lancet. 2021;398(10304):944–5.

